# Correlates of reproductive coercion among young women in Kingston, Jamaica and Hanoi, Vietnam

**DOI:** 10.1101/2025.05.23.25328228

**Authors:** June Postalakis, Tina Hylton-Kong, Nghia Nguyen, Eliana Burlotos, Alison Norris, Maria F. Gallo

## Abstract

**Objectives:** We sought to identify the correlates of reproductive coercion (RC) among women in Kingston, Jamaica and Hanoi, Vietnam.

**Study design:** We analyzed data from two cross-sectional studies: 1) a study of 222 women, 18-25 years of age, attending a clinic in Kingston, Jamaica in 2018-2019 and 2) a study of 500 women, 18-45 years of age, receiving care at a hospital in Hanoi, Vietnam in 2017-2018. Shared eligibility criteria between the populations included being sexually active and not desiring pregnancy. We categorized women as experiencing RC if they indicated that their male partner had engaged in at least one of the following: had pressured them to become pregnant, would stop them if they wanted to use a method to prevent pregnancy, had messed with or made it difficult to use a method to prevent pregnancy, or had ever stopped them from using a method to prevent pregnancy. We used logistic regression to examine associations between demographic factors and experiencing RC.

**Results:** RC from a male partner was common in both populations, with a prevalence of 44% in Jamaica and 17% in Vietnam. Lower educational attainment was correlated with RC in Jamaica. Correlates in Vietnam included younger age and a history of forced sex or experiencing intimate partner violence.

**Conclusions:** RC among women in Jamaica and Vietnam appeared to be more common among those who held other characteristics associated with low power. Considering the role of a male partner is critical to promote reproductive justice.

**Implications:** Reproductive coercion from a male partner was more common with lower educational attainment in Kingston, Jamaica and among women of younger age and a history of forced sex or intimate partner violence in Hanoi, Vietnam.

## 1. Introduction

Reproductive autonomy, defined as the ability to make decisions about and control one’s behaviors related to fertility and contraception use, is vital for people’s health and well-being [1]. Reproductive autonomy can promote contraceptive uptake [2]; conversely, reproductive coercion (RC) can pose a barrier to contraception use or result in someone becoming pregnant unwillingly [3-5]. Although various individuals, including in-laws [6], can exert RC, male partners often are a primary perpetrator.

Male partners exert RC for various reasons, including desire for children; misconceptions about contraceptive side effects and the belief that they can lead to women’s infidelity; and reduced sexual pleasure from contraceptive use [3, 7-9]. RC often manifests as instructing a female partner not to use contraception, forcing her to become pregnant, or engaging in contraceptive sabotage (e.g., damaging condoms or removing them mid-act or destroying or hiding pills) [5]. Less direct forms occur when women anticipate or perceive their partner disapproval of contraceptive use [9,10]. Experiencing or anticipating RC can cause feelings of distress, anger, and trauma [11,12]. Women may adopt various strategies to deal with perceived or encountered resistance, such as “sweet talk,” referencing experts, eroticizing condom use [11,13] or using contraception covertly. The latter can be anxiety producing and challenging logistically, especially when women are financially dependent on their partner [14]. Intimate partner violence (IPV) can increase the risk of RC [15], and women may hesitate to voice desire for condom use because of past experiences with verbal arguments or physical violence [13]. Healthcare providers who anticipate male resistance may be reluctant to offer contraceptive counseling, thus compounding barriers to contraception access [14]. RC is associated with increased care seeking for pregnancy or sexually transmitted infection testing and emergency contraception [16] and could contribute to poor mental health, including an increased risk of depression and post-traumatic stress disorder [17,18]. Contraceptive sabotage can compromise women’s sexual health or lead to unintended pregnancy [19].

We sought to identify correlates of RC among women in two settings: Kingston, Jamaica and Hanoi, Vietnam. Cultural norms in Jamaica could complicate women’s ability to negotiate condom use [13]. Masculinity and inequitable gender norms have been associated with Jamaican men’s desire for high fertility and a reduced focus on preventing unplanned pregnancy [20]. In Vietnam, traditional gender roles remain common, with most men playing a leading role in the family-marriage relationship [21]. Additionally, patriarchal ideas rooted in Confucianism can influence reproductive decision making and family planning [22].

## 2. Materials and methods

### 2.1 Jamaica study population

We analyzed enrollment data from a randomized controlled trial (N=225) conducted at a large public clinic in Kingston, Jamaica from November 7, 2018 to March 28, 2019. Young (18-25 years of age) women who were sexually active and not desiring pregnancy within the next year were eligible. Those using long-acting contraception, pregnant, or breastfeeding were excluded. The study, designed to evaluate the effect of a video intervention on beliefs about intrauterine devices and implants, has been described elsewhere [23]. Female study interviewers administered the enrollment questionnaire, which included questions on demographics, contraceptive use, sexual activity, reproductive history, and contraceptive-related knowledge. Only women who provided written consent were enrolled, and the Ohio State University Institutional Review Board (OSU IRB) and the Jamaica Ministry of Health Ethics Committee provided ethical review. Retrospective data was accessed for research purposes on September 27, 2022 and authors did not have access to information that could identify individual participants.

### 2.2 Vietnam study population

We analyzed data from a cross-sectional survey (N=500) conducted in the obstetrics-gynecology department of a large public hospital in Hanoi, Vietnam from November 1, 2017 to September 30, 2018. Participants were of reproductive age (18-45 years), sexually-active, and not desiring pregnancy in the next year. Similar to the Jamaica population, women who were pregnant or breastfeeding were excluded, as well as those without a minimal level of literacy or comfort using a computer. The study was designed to test an intervention to improve contraceptive knowledge and has been described elsewhere [24]. Female study staff administered a questionnaire containing questions regarding demographics, sexual activity, reproductive and contraception use history, IPV, and contraceptive-related knowledge and attitudes. Only women who provided written consent were enrolled, and the OSU IRB and the Hanoi School of Public Health IRB approved the research. Retrospective data was accessed for research purposes on September 27, 2022 and authors did not have access to information that could identify individual participants.

### 2.3 Reproductive coercion measure

The main outcome of the present analysis was women’s reports of RC. We assessed RC using the following four items: “My partner has pressured me to become pregnant,” “If I wanted to use a method to prevent pregnancy my partner would stop me,” “My partner has messed with or made it difficult to use a method to prevent pregnancy when I wanted to use one,” and “My partner has stopped me from using a method to prevent pregnancy when I wanted to use one.” We collapsed the Likert scale response option into two categories: “strongly agree” or “agree” vs. “disagree” or “strongly disagree.” We categorized women as experiencing RC if they agreed with at least one item. We found acceptable internal consistency for the derived outcome measure for both populations (Cronbach’s alpha of 0.66 and 0.63 for the Jamaica and Vietnam datasets, respectively).

### 2.4 Correlates in Jamaica study

We assessed the following potential correlates of RC: age (separated into terciles of 18-19, 20-22, 23-25 years), education completed (high school or less vs. vocational skills/training vs. some college/university), union status (married/common-law/cohabiting vs. other), current employment (part-time or full-time job vs. neither), currently in school (yes vs. no), live children (yes vs. no), currently using a method to prevent pregnancy use (yes vs. no), most effective method by male/female control (not using contraception vs. male vs. female control), and partner knowledge of contraceptive method (yes and not using a condom vs. yes and using a condom vs. no). To assess control of contraceptive method, we first categorized women by their most effective method and then categorized method use as under male control (i.e., male condom, fertility-based awareness, withdrawal) or female control (i.e., oral contraception and injectable contraception).

### 2.5 Correlates in Vietnam study

We assessed the following possible correlates of reported RC: age (21-31, 32-36, 37-45 years), residence (urban vs. town or rural area), education completed (upper secondary or less vs. higher), currently in school (yes vs. no), at least weekly sexual frequency (yes vs. no), current contraception use (male control vs. female control using the categorization described above), partner aware of method use (yes vs. no), live children (yes vs. no), history of being forced or pressured to have sex (yes vs. no), history of physical IPV (yes vs. no, measured using an established set of questions [25]), and sexual autonomy (yes vs. no). Lacking “sexual autonomy” was based on responding “no” to at least one of the following questions, “In your opinion, can a married woman refuse to have sex with her husband if:” (1) “she doesn’t want to,” (2) “he is drunk,” (3) “she is sick,” or (4) “he mistreats her?”

### 2.6 Statistical analysis

For Jamaica and Vietnam, separately, we conducted univariate logistic regression to identify possible correlates of reporting RC and then created an adjusted model based on factors that have been associated with RC in the literature: age, urban residence, education, union status, and having live children [3,4,6,15,26-30]. We could not adjust for urban residence in Jamaica as this variable was not collected and could not adjust for union status or live children in Vietnam because of insufficient diversity.

## 3. Results

### 3.1 Prevalence of RC

About 44% of women in Jamaica reported experiencing at least one type of RC, compared to 17% of women in Vietnam (Table 1). The most commonly reported type in Jamaica was that their partner would stop them if they wanted to use a method to prevent pregnancy (24%). In Vietnam, the most commonly reported type was that their partner has stopped then from using a method to prevent pregnancy when they wanted to use one (13%).

**Table 1.** Prevalence of reproductive coercion among women in Jamaica, 2018-2019 (N=222) and Vietnam, 2017-2018 (N=500)

|  | Jamaica<br>N (%) | Vietnam<br>N (%) |
| --- | --- | --- |
| If I wanted to use a method to prevent pregnancy my partner would stop me. | 54 (24.4) | 41 (8.2) |
| My partner has pressured me to become pregnant. | 45 (20.4) | 7 (1.4) |
| My partner has messed with or made it difficult to use a method to prevent pregnancy when I wanted to use one. | 46 (20.7) | 28 (5.6) |
| My partner has stopped me from using a method to prevent pregnancy when I wanted to use one. | 37 (16.7) | 64 (12.8) |
| One or more of these | 98 (44.1) | 86 (17.2) |

### 3.2 Correlates of RC in Jamaica

The mean age in Jamaica was 21 years, with a standard deviation of 2.4 (Table 2). Most women surveyed had a high school education or less (57%) and did not have a job (62%). One variable was statistically significantly associated with reporting RC in the unadjusted analysis for the Jamaica dataset (Table 3). Women who completed high school or less had 2.8 times greater odds of reporting RC compared to women with more education (95% CI: 1.2–6.5). In the adjusted analysis, the odds of reporting RC remained higher among women with a high school educational level or less compared to women with more education (AOR 3.1, 95% CI: 1.3–7.5). We did not find an association between other variables and reporting RC.

**Table 2.** Reproductive coercion by participant characteristics among sexually active women in Kingston, Jamaica, 2018-2019 (N=222)

|  | Reproductive coercion |  |
| --- | --- | --- |
|  | Yes (n=98)<br>n (%) | No (n=124)<br>n (%) |
| Age (years) |  |  |
| 18-19 | 28 (28.6) | 42 (33.9) |
| 20-22 | 39 (39.8) | 33 (26.6) |
| 23-25 | 31 (31.6) | 49 (39.5) |
| Education completed |  |  |
| High school or less | 64 (65.3) | 63 (51.2) |
| Vocational skills/training | 25 (25.5) | 35 (28.5) |
| Some college/university | 9 (9.2) | 25 (20.3) |
| Union status |  |  |
| Married, common-law, cohabiting | 30 (30.6) | 47 (38.5) |
| Other | 68 (69.4) | 75 (61.8) |
| Currently employed (part-time or full-time) |  |  |
| Yes | 37 (37.8) | 47 (37.9) |
| No | 61 (62.2) | 77 (62.1) |
| Currently in school |  |  |
| Yes | 24 (24.5) | 41 (33.6) |
| No | 74 (75.5) | 81 (66.4) |
| Live children |  |  |
| Yes | 44 (46.3) | 56 (45.2) |
| No | 51 (53.7) | 68 (54.8) |
| Most effective method currently using to prevent pregnancy |  |  |
| Injectable | 15 (15.6) | 20 (16.7) |
| Oral contraception | 4 (4.2) | 6 (5.0) |
| Male condom | 26 (27.1) | 42 (35.0) |
| Fertility-based awareness, withdrawal | 0 (0.0) | 2 (1.7) |
| Not using contraception | 51 (53.1) | 50 (41.7) |
| Partner aware of contraceptive method use |  |  |
| Yes, not using condom | 39 (41.9) | 44 (37.9) |
| Yes, using condom | 37 (39.8) | 56 (48.3) |
| No or unsure | 17 (18.3) | 16 (13.8) |

The following questions had missing responses: union status (2), educational level (1), educational status (2), live children (3), partner knowledge of contraceptive methods (13).

**Table 3.** Correlates of reporting reproductive coercion among sexually active women in Kingston, Jamaica, 2018-2019 (N=222)

|  | OR (95% CI) | Adjusted OR (95% CI)* |
| --- | --- | --- |
| Age (years) |  |  |
| 18-19 | 1.1 (0.5, 2.0) | 0.9 (0.4, 1.8) |
| 20-22 | 1.9 (0.9, 3.6) | 1.7 (0.9, 3.3) |
| 23-25 (ref) | - | - |
| Education completed |  |  |
| High school or less | 2.8 (1.2, 6.5) | 3.1 (1.3, 7.5) |
| Vocational skills/training | 2.0 (0.8, 5.0) | 2.0 (0.8, 5.3) |
| Some college/university (ref) | - | - |
| Union status |  |  |
| Married, common-law, cohabiting | 0.7 (0.4, 1.2) | 0.6 (0.3, 1.2) |
| Other (ref) | - | - |
| Currently employed (part-time or full-time) |  |  |
| Yes | 1.0 (0.6, 1.7) | 1.0 (0.6, 1.9) |
| No (ref) | - | - |
| Currently in school |  |  |
| No | 1.6 (0.9, 2.8) | 1.4 (0.7, 2.8) |
| Yes (ref) | - | - |
| Live children |  |  |
| Yes | 1.0 (0.6, 1.8) | 1.1 (0.6, 2.1) |
| No (ref) | - | - |
| Currently using a method to prevent pregnancy |  |  |
| No | 1.6 (0.9, 2.7) | 1.5 (0.9, 2.7) |
| Yes (ref) | - | - |
| Most effective method by male/female control |  |  |
| Not using contraception | 1.4 (0.7, 2.8) | 1.3 (0.6, 2.7) |
| Male control | 0.8 (0.4, 1.7) | 0.7 (0.3, 1.7) |
| Female control (ref) | - | - |
| Partner aware of contraceptive method use |  |  |
| No or unsure | 1.2 (0.5, 2.7) | 1.1 (0.5, 2.5) |
| Yes, using condom | 0.7 (0.4, 1.4) | 0.8 (0.4, 1.5) |
| Yes, not using condom (ref) | - | - |

OR = odds ratio; CI = confidence interval
\*Adjusted for age, education completed, live children, and union status.

### 3.3 Correlates of RC in Vietnam

The mean age of women surveyed in Vietnam was 34 years, with a standard deviation of 5.4 (Table 4). Almost all women surveyed resided in an urban area (90.0%), were married or cohabitating (97.2%), were currently working (93.0%), and had live children (95.0%). Three variables were statistically significantly associated with reporting RC in the unadjusted analysis for the Vietnam dataset (Table 5). Women aged 21-31 years had greater odds of reporting RC compared to older women (OR 1.9, 95% CI 1.0–3.3). Women who had a history of forced sex (OR 2.0, 95% CI 1.0–3.8) or physical IPV (OR 2.3, 95% CI 1.4–3.9) had greater odds of reporting RC. Variables remained statistically significant when adjusting for age, union, and residence.

**Table 4.** Reproductive coercion by participant characteristics among sexually active women in Hanoi, Vietnam, 2017-2018 (N=500)

|  | Reproductive coercion |  |
| --- | --- | --- |
|  | Yes (n=86)<br>n (%) | No (n=414)<br>n (%) |
| Age (years) |  |  |
| 21-31 | 35 (40.7) | 121 (29.2) |
| 32-36 | 28 (32.6) | 145 (35.0) |
| 37-45 | 23 (26.7) | 148 (35.7) |
| Education completed |  |  |
| Upper secondary or less | 27 (31.4) | 100 (24.2) |
| Higher | 59 (68.6) | 313 (75.8) |
| Union status |  |  |
| Married or cohabiting | 84 (97.7) | 402 (97.3) |
| Not married or cohabiting | 2 (2.3) | 11 (2.7) |
| Residence |  |  |
| Urban area | 73 (84.9) | 377 (91.1) |
| Town or rural area | 13 (15.1) | 37 (8.9) |
| Currently employed |  |  |
| Yes | 80 (95.2) | 385 (93.2) |
| No | 4 (4.8) | 28 (6.8) |
| Currently in school |  |  |
| Yes | 11 (12.8) | 54 (13.1) |
| No | 75 (87.2) | 358 (86.9) |
| Sexual frequency |  |  |
| At least once per week | 61 (78.2) | 347 (86.3) |
| Less than once per week but at least every other week or monthly | 17 (21.8) | 55 (13.7) |
| Most effective method currently using to prevent pregnancy |  |  |
| Male condom | 31 (36.5) | 135 (32.8) |
| Intrauterine device | 18 (21.2) | 108 (26.2) |
| Oral contraception | 18 (21.2) | 106 (25.7) |
| Fertility-based awareness, withdrawal, other | 9 (10.6) | 49 (11.9) |
| Female condom | 2 (2.4) | 5 (1.2) |
| Implant | 2 (2.4) | 3 (0.7) |
| Injectable | 0 (0.0) | 1 (0.2) |
| Male partner sterilization | 1 (0.0) | 0 (0.0) |
| Not using contraception | 4 (0.0) | 5 (1.2) |
| Partner is aware of contraceptive method use |  |  |
| Yes | 81 (100.0) | 406 (99.8) |
| No | 0 (0.0) | 1 (0.2) |
| Live children |  |  |
| Yes | 80 (94.1) | 395 (96.3) |
| No | 5 (5.9) | 15 (3.7) |
| History of forced sex |  |  |
| Yes | 15 (17.4) | 39 (9.5) |
| No | 71 (82.6) | 370 (90.5) |
| History of physical intimate partner violence |  |  |
| Yes | 26 (31.3) | 68 (16.6) |
| No | 57 (68.7) | 342 (83.4) |
| Sexual autonomy |  |  |
| Yes | 74 (90.2) | 369 (93.2) |
| No | 8 (9.8) | 27 (6.8) |

The following questions had missing responses: union status (1), educational level (1), currently in school (2), currently employed (3), sexual frequency (20), current contraceptive method (13), live children (5), partner aware of contraceptive method use (12), history of forced sex (5), history of physical intimate partner violence (7), and sexual autonomy (22).

**Table 5.** Correlates of reporting reproductive coercion among sexually active women in Hanoi, Vietnam, 2017-2018 (N=500)

|  | OR (95% CI) | Adjusted OR (95% CI)* |
| --- | --- | --- |
| Age (years) |  |  |
| 21-31 | 1.9 (1.0, 3.3) | 1.9 (1.0, 3.3) |
| 32-36 | 1.2 (0.7, 2.3) | 1.2 (0.7, 2.2) |
| 37-45 (ref) | - | - |
| Residence |  |  |
| Town or rural area | 1.8 (0.9, 3.6) | 1.5 (0.7, 3.1) |
| Urban area (ref) | - | - |
| Highest level of education completed |  |  |
| Upper secondary or less | 1.4 (0.9, 2.4) | 1.4 (0.8, 2.3) |
| Higher (ref) | - | - |
| Currently in school |  |  |
| No | 1.0 (0.5, 2.0) | 0.9 (0.5, 1.9) |
| Yes (ref) | - | - |
| At least weekly sexual frequency |  |  |
| No | 1.8 (1.0, 3.2) | 1.7 (0.9, 3.2) |
| Yes (ref) | - | - |
| Current contraception method |  |  |
| Male control method | 1.2 (0.7, 2.0) | 1.2 (0.7, 1.9) |
| Female control method (ref) | - | - |
| History of forced sex |  |  |
| Yes | 2.0 (1.0, 3.8) | 2.0 (1.0, 3.8) |
| No (ref) | - | - |
| History of physical intimate partner violence |  |  |
| Yes | 2.3 (1.4, 3.9) | 2.2 (1.3, 3.9) |
| No (ref) | - | - |
| Sexual autonomy |  |  |
| Yes | 1.5 (0.6, 3.3) | 1.4 (0.6, 3.3) |
| No (ref) | - | - |

OR = odds ratio; CI = confidence interval
\*Adjusted for age, rural/urban residence, and education level.

## 4. Discussion

Reports of experiencing RC were common among young women in Jamaica, with 44% reporting at least one type. Women with a low educational level were particularly vulnerable. Reporting RC was less common among the women studied in Vietnam but still a pervasive issue, with almost one fifth reporting at least one type. Younger women and those with a history of physical IPV or forced sex had higher odds of reporting RC. This has important implications for the sexual health of women and their agency.

The most common type of RC reported in Jamaica was a belief: anticipation that her partner would stop her if she tried to use contraception. This aligns with evidence in the literature demonstrating that perceived and anticipated resistance can prevent contraceptive uptake [9,10] and illustrates that RC in Jamaica may be mediated by inequitable gender norms [20]. In contrast, the most common type of RC reported in Vietnam was an experience: a partner stopping a woman from using contraception. This may be due to men’s dominant role in reproductive decision-making in Vietnam, as men often decide on the contraceptive method to be used [31,32] and their control over method use may be widely perceived as acceptable. It also may be related to the greater proportion of women in Vietnam who were married or cohabiting.

While the link between RC and influence on contraceptive uptake has been demonstrated, few studies have examined correlates of RC. Limited evidence suggests that younger women and women cohabiting with in-laws may face increased risk [3,6]. Other correlates include living in a rural area, having experienced IPV, being single or dating, being black or multiracial, lacking health insurance, and lower household wealth [6,15,19,26,29,33]. Higher levels of education and parity can be protective against RC [26,27]. The correlates identified in this study all align with existing literature. Younger age was associated with RC in the Vietnam population but was not a statistically significant variable in the Jamaica population, perhaps because it was a young population. Educational level was significant in the Jamaica population rather than in the Vietnam population, which could similarly be due to the different age ranges of participants. Several correlates identified in the literature (living in a rural area, parity, and being single or dating) were not significantly associated with RC in this study. Lack of consistency in the identification of correlates between studies could be due to true differences in population characteristics or could reflect methodological issues related to low study power or lack of heterogeneity in the study samples for some of the attributes which could have prevented the ability to detect relationships.

A strength of the present study was the use of the same four items for measuring RC in two distinct populations. The items had good internal consistency, which supported their combination into a single measure. Limitations included the convenience sampling method and cross-sectional design. The Jamaica analysis was limited by its small sample size while the Vietnam sample was limited by its homogenous nature, with little variation in key demographic variables. Both analyses were limited by the potential for social desirability bias and misclassification of women who experienced partner pressure to become pregnant with a healthy and supportive sexual partner. There were also several missing responses for variables in both populations, including partner knowledge of contraceptive methods in Jamaica, and sexual frequency and sexual autonomy in Vietnam. Additionally, because women can also be unaware of some forms of RC, such as condom removal during sex, the measures might be underestimations.

Male partners can greatly influence their female partner’s access to contraception, which can endanger women’s mental, physical, and emotional well-being. Interventions to identify women are at risk of RC are needed to be able to prevent pregnancy coercion and support women in leaving an unhealthy or unsafe relationship [34,35]. Measuring RC is critical for raising awareness and changing societal norms about the value of reproductive autonomy. Future research should collect data from dyads to better inform contraceptive counseling and intervention development. To improve contraceptive access, healthcare providers must consider the role of the partner in decision-making to best support and empower women. Not only does RC curtail contraceptive access, but it also violates human rights by infringing on bodily autonomy. Efforts to promote reproductive justice and human rights should consider RC as a significant but modifiable barrier.

## Data Availability

N/A

## Acknowledgements

This work was supported by the Bill & Melinda Gates Foundation (OPP1171894), the Society of Family Planning Research Fund (SFPRF11-04), and the National Center for Advancing Translational Sciences (UL1TR001070). The sponsors had no role in the study design, collection, analysis and interpretation of data, writing of the report, and decision to submit the article for publication.

